# Covid-19 in Brazil: A Cross-Sectional Immuno-Epidemical and Genomic Evaluation in a Pre-Omicron Era

**DOI:** 10.1101/2022.11.12.22282248

**Authors:** Sarah de Oliveira Rodrigues, Gabriel Ferrari de Oliveira, Marcella Pâmela Costa e Silva, Jorge Pamplona Pagnossa

## Abstract

The seventh human coronavirus, was discovered and reported primarily in Wuhan, China. After intense seasons with repercussions in all areas ofhumanity, the pandemic demonstrates a new perspective. In Brazil, the pandemic concept has had impacts in vast areas, including mainly hospitals. This present study aims to describe, present and synthesize data that correlate the symptoms of passive and/or active patients for Covid-19 and their respective results of igg/igm serological tests in hospitals in cruzeiro, São Paulo. The form had been applied to 333 people and obtained conclusive results and several symptoms presented, in addition, asymptomatic cases were also analyzed and directed in the genomic study of variants of concern, as well as vaccination data in the study region.

## 1. INTRODUCTION

In 2019, an outbreak of pneumonia of unknown origin affected citizens in wuhan city, Hubei province, China. Subsequently, the viral infection spread in an aritmetic way around the world, being declared a pandemic by the WHO Director General in 2020 (1). Like other respiratory viruses, transmission of SARS-CoV-2 occurred with high efficiency and ctivityinfection, mainly through the respiratory tract. Droplet transmission is the main known route, although aerosols can also be highlighted (2).

Several types of tests were used to detect the immunological response against Covid-19 in humans, including IgM/IgG rapid diagnosis tests performed at the time of hospital admission. The body produces two main types of antibodies, immunoglobulin M (IgM) and immunoglobulin G (IgG), in response to an infection. IgM antibodies are produced soon after infection, while IgG antibodies are subsequently produced to maintain the immune system with the necessary genetic information (3).

At the time of admission of the patient to the hospital unit, a pre-selection of screening is performed, in this intermediary, a questionnaire about symptomatic signs had been applied in an inclusive way, seeking reliable results on the serological tests of rapid diagnosis, as well as the main symptoms presented by the paci entities withinthe pandemic context.

Moreover, the common genetic components of the Variants of SARS-CoV-2 were also evaluated and characterized in order to directly estimate the adoption of hygienic-sanitary and/or medicinal measures of an experimental character in a hospital and community environment, since the lack of engagement of the population to effective measures against the spread of the virus results in the emergence of new variants, possibly more infectious and lethal.

## 2. METHODOLOGY

The present study occurred in the pandemic period described between June and November 2021 with the application of a simple model questionnaire, covering age, gender and vaccination as general classifiers and 20 symptoms that could be marked facultatively between “yes”, “no” or “do not know”. In addition to this questionnaire, serological tests were performed for the presence of IgG and IgM antibodies as a way to correlate the symptoms and immunological response of each individual. The symptoms mentioned in theperiod mentioned were the same as those evidenced by the World Health Organization in the specific period described. Among the reported symptoms are: fever, sore throat, rinorrhoea, cough, shortness of breath, chills, vomiting, nausea, diarrhea, headache, coceira, conjunctivitis, myalgia, joint pain, loss of appetite, loss of smell, loss of taste, nasal bleeding, fatigue and convulsions.

Directly correlated with the IgG and IgM results of the serological tests performed, such data were collected, organized and synthesized in specific didactic formats (graphs and tables). In addition, the present study performedcareful bibliographic aspects of symptom analysis, social impacts, historical context and general theoretical basis in the pandemic context, using research sources such as PubMed, Scielo and Google Scholar.

For genomic evaluation, bioinformatics tools are used. In this stage of the research, computational resources such as Python algorithms (developed by the authors themselves) and the Nextclade application were used. The first Python program was responsible for collecting *from the National Center for Biotechnology Information* (NCBI) database the nucleotide sequences of SARS-CoV-2 samples for the period and country previously specified. The genomes were then inserted into the Nextclade software that compared them to the genome of the original Wuhan strain and then provides the information regarding the mutations that occurred. The second *Python script* was responsible for processing the information generated by Nextclade.

The keywords highlighted in this analytical and descriptive epidemiological research are: “Covid-19”, “pandemic”, “nalysis”, “population”, “hospitals” and “SARS-CoV-2”, used without exclusion criteria. Thus, articles that were in the pre-established period had greater attention and relevance for the greater applicability in the study.

## 3. RESULTS AND DISCUSSION

### 3.1 FREQUENT SYMPTOMS

In a contextual parameter, the global crisis caused by the virus permeates society more devastatingly in developing countries due to the precariousness of hospitals and health centers. As of April 2021, there were more than 140,000confirmed cases and more than three million deaths from Covid-19 worldwide (4). Although vaccination began in several countries, new outbreaks and variants continued to emerge. Similarly, the global distribution of vaccines is hampered by equity challenges, as well as logistical complications.

Moreover, in the Brazilian context, the pandemic became a major challenge for hospital institutions in general, vaccination was still incipient and knowledge about the resolution of symptoms was still being implemented. However, certain symptomatic patterns were characteristic izedby the WHO, evidencing diagnoses and providing greater margin of positive predictive. In view of this, the collection of data in the hospital scope that had been performed between June and November 2021 demonstrates the frequency of symptoms in active or passive paci entities forviral immune response, which sought care in health institutions in specific regions of the city of Cruzeiro-SP.

From the survey of the symptoms of people, with or without positivity for immunological response due to Covid-19 (Figure 1), respiratory tract involvement is the most prevalent (cough: 60.0%; and rinorrhoea: 41.67%), accompanied by cefaleia (55.83%) and skeletal muscle disorders (fatigue: 37.50; myalgia: 36.67%; and joint pain: 30.0%) and gastrointestinal (loss of appetite: 28.33%; and diarrhea: 25.0%). During the occurrence of a pandemic that has been the case with influenza-likeviral origin, the diversity of symptoms is a key factor in the patient’s decision to seek care at health centers in order to perform serological screening tests, or even to confirm the presence of the virus in the oropharine tract.

**Figure 1.**
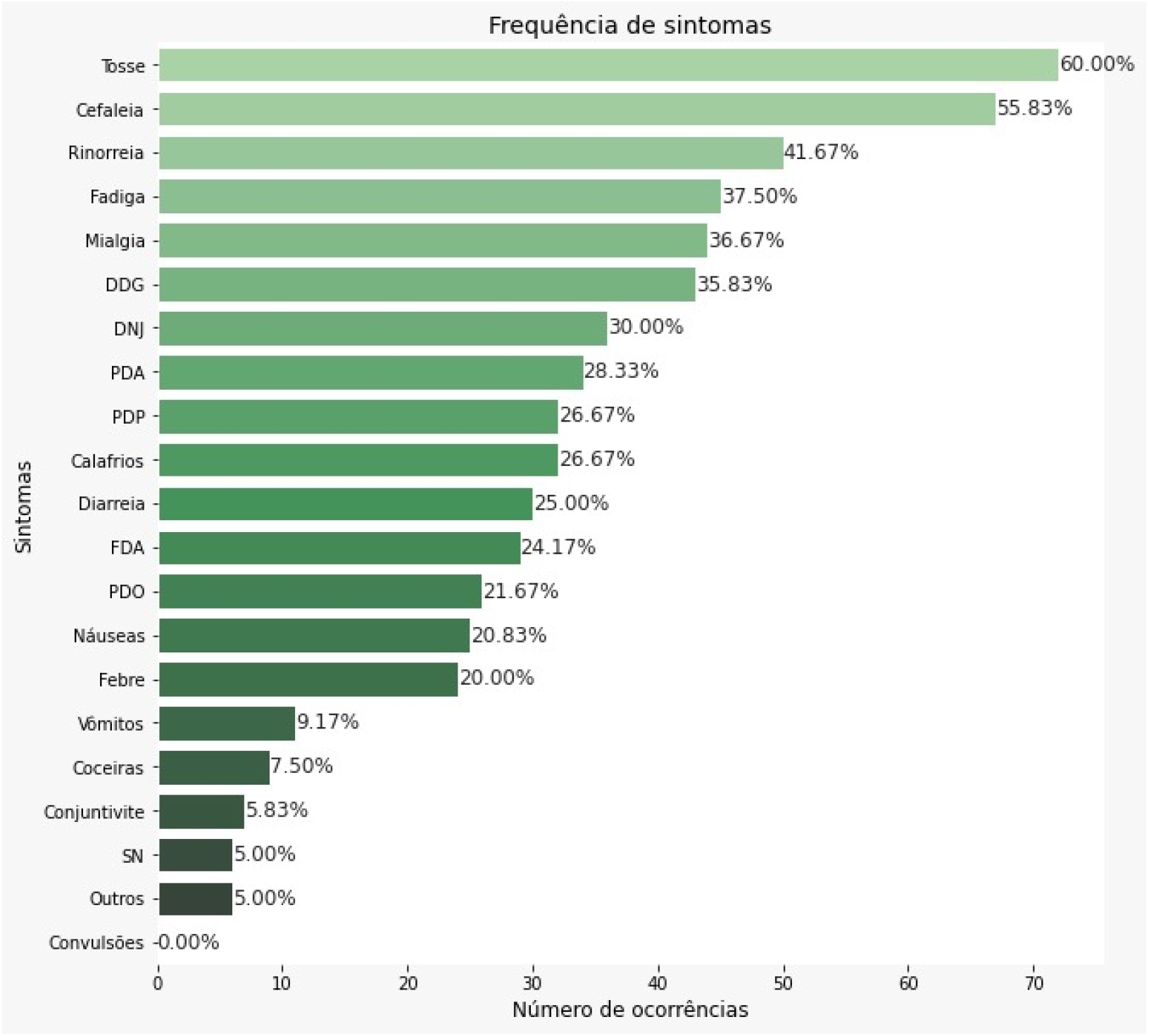
Frequency of symptoms in research participants. Source: Developed by the Authors (2022)

The frequency of symptoms was correlated with the predictive symptoms most evident in the literature. Recent studies have demonstrated the involvement of the naso-oropharyngeal tract as a possible typical class of symptoms to diagnose cases of Covid-19, primarily evidencing the occurrence of cough (43.8%) (5)

In the anatomical sphere, it is clear that the structures affected by the most common symptoms have full communication with each other (figure 2). Cough (a natural reflex of the respiratory system that arises as a consequence of an irritative or inflammatory process) (6), and the rinorreia (excessive discharge of nasal mucus resulting from infectious or allergic process) (7), are constantly related to bacterial or viral infections, which affect the respiratory system as a whole.

**Figure 2.**
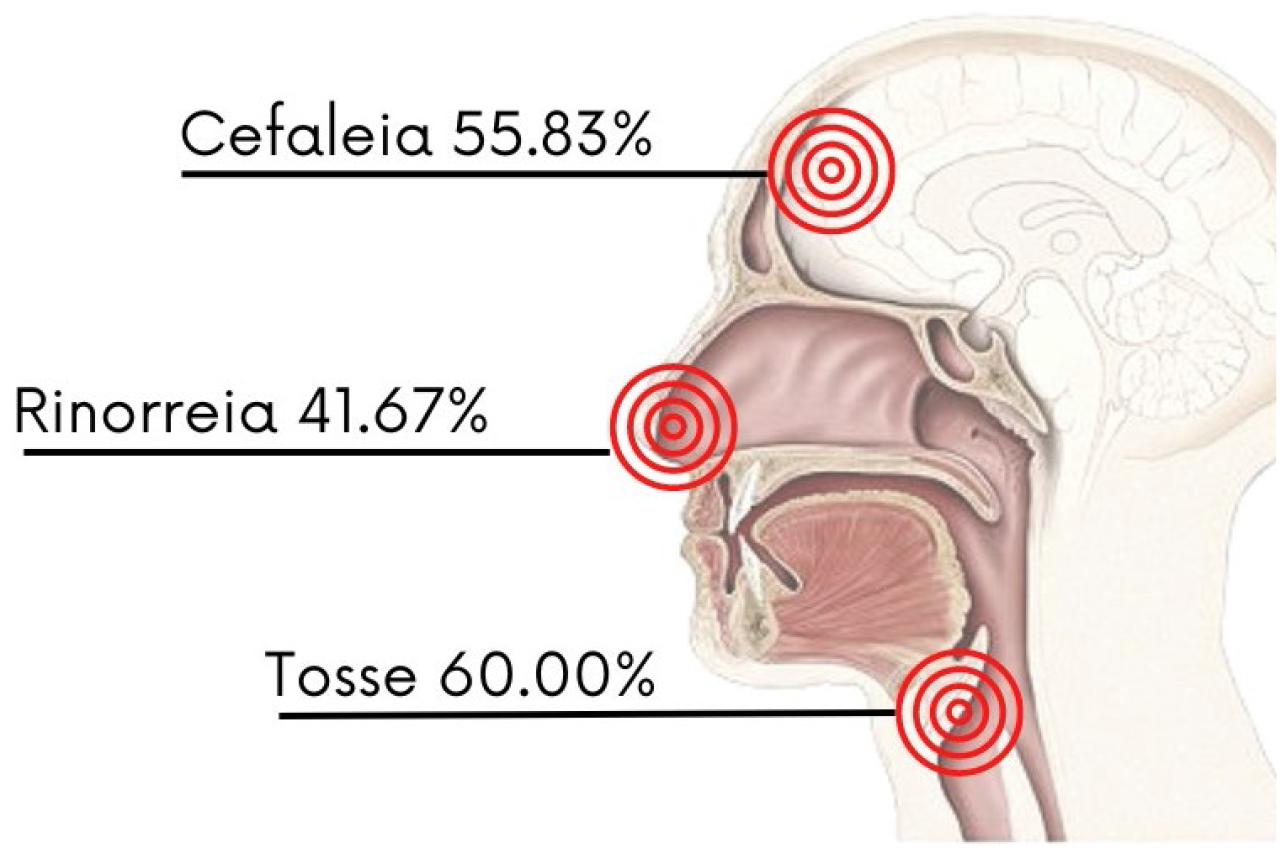
Representation of the anatomical proximity of the areas affected by symptoms: headache, rinorrea and cough. Source: Developed by the Authors (2022)

After identification of the microrganism, one of the cleaning systems by secretion of liquids of the organism is activated, located in the part of the cavities of the skull (8). The mucous wall present in the paranasal sinus has constituents with immunological properties, nasal secretions contain immunoglobulins IgA, IgG, IgM, IgE, and enzymes such as lysozymes and lactoferrin, protective proteins as a complement and also neutrophils and lymphocytes in the mucosa (9), fully supporting in infectious processes. However, the accumulation of mucus in the paranasal sinus can cause primarily headache (feeling of full pain in the head region). In short, the symptoms of cough, rinorreia and headache are fully intertwined in immunological processes, as well as in the fight against the SARS-CoV-2 virus.

Thus, it is emphasized that the symptomatology obtained different classifications, as well as the number of symptoms presented by each patient. After data processing, it was possible to observe (Figure 3) that 21.7% of the people presented only one, two or three distinct symptoms and in greater quantidade, 29.2% of the patients had four, five or six symptoms, evidencing that the symptoms affected by the virus are mostly multiple. In addition, the number of symptoms between seven and twelve represent 28.3%, emphasizing the concept of symptomatic somatium, which is based on primordial characteristic symptoms, which culminate in other symptoms of similar origins, physiologically and anatomically (10).

**Figure 3.**
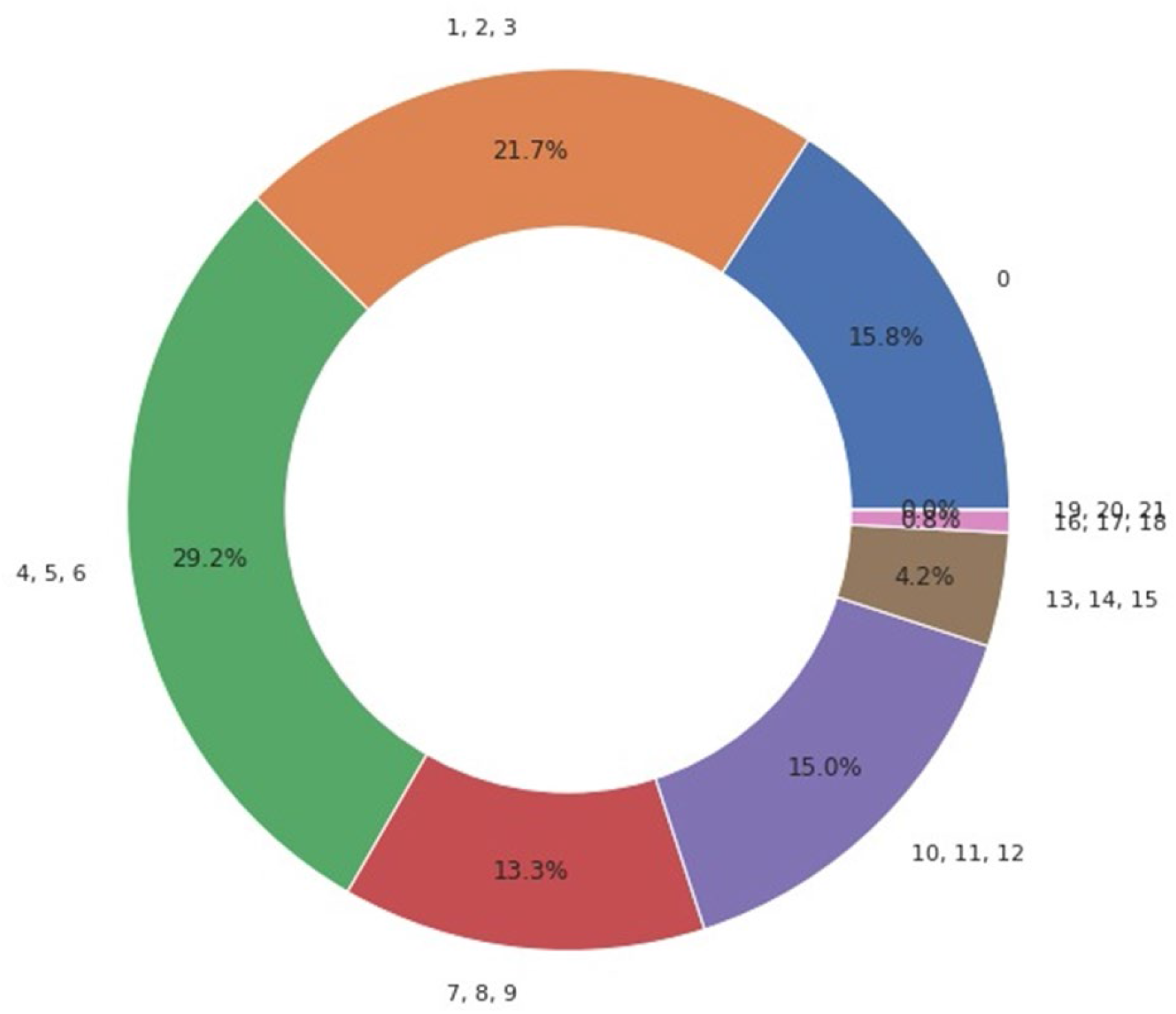
Number of symptoms per person. Source: Developed by the Authors (2022)

In addition, the number of symptoms per person has direct proportionality with the number of patients present in the study. A fundamental numerical characteristic for the analysis of the group as a whole is the age factor (Figure 4).

**Figure 4.**
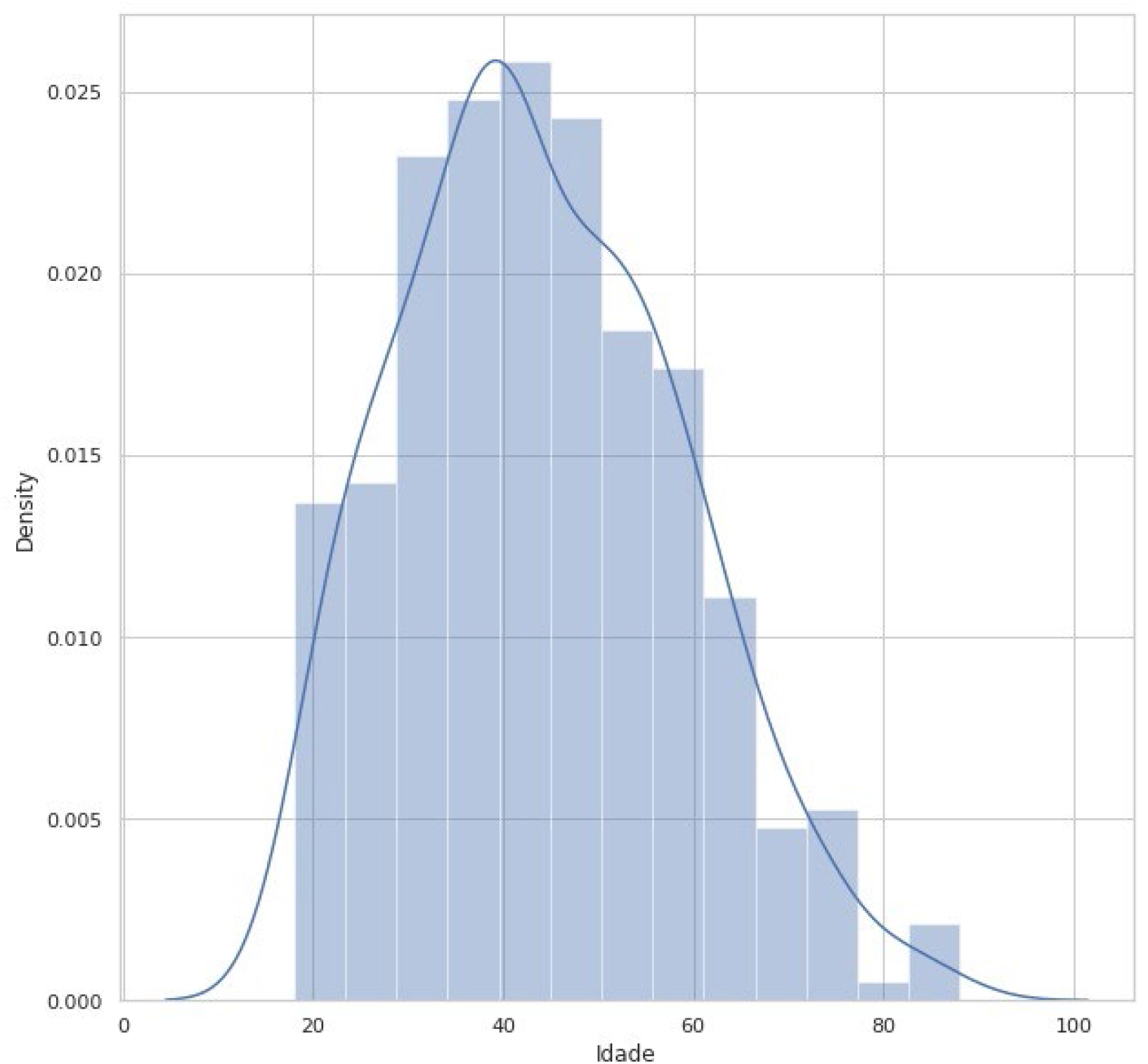
age group by population density. Source: Developed by the Authors (2022)

During the specific period of time that the study was conducted, informative perspectives were implemented in the city of Cruzeiro-SP, decreed by the state of São Paulo, with prescription of who guidelines (11). The instructions were to guide the population on hospital safety norms, being certain specific groups of higher priority in basic health units, seeking to avoid stocking and greater probability of contamination and spread of the virus (12), both for patients and non-patients (critical andpost-diagnosis). However, the visible highlight in age, obtained through the data collected between July and November 2021, has its highest mean age between 35 and 60 years, evidencing 45 years specifically (Figure 4), as the mostpart-time age group in the study. Demonstrating the correlation between age and prioritization of care for cases of specific or severe symptoms.

The public participatedbefore the study of up to 60 years of age is among the highest percentage of age because it was one of the current recommendations to seek care and diagnosis from the symptomatic involvement, later, the patient was led to medical evaluation, after the procedure, serological tests were performed, in large majority, only cases with probable severity were prioritized, excluding adolescents and children in the risk group. Data from serological tests correlated with the proportionality of symptoms reported by patients (Figure 5) were also processed, showing a higher frequency of negative diagnoses per symptom, i.e., although participants reported symptoms such as fatigue, diarrhea and fever, negative results in immunological tests indicate the possibility of involvement by flu-like syndromes similar to Covid-19. Clearly, symptoms are characteristic of other diseases, such as inflammation of the gastrointestinal tract of bacterial origin, for example.

**Figure 5.**
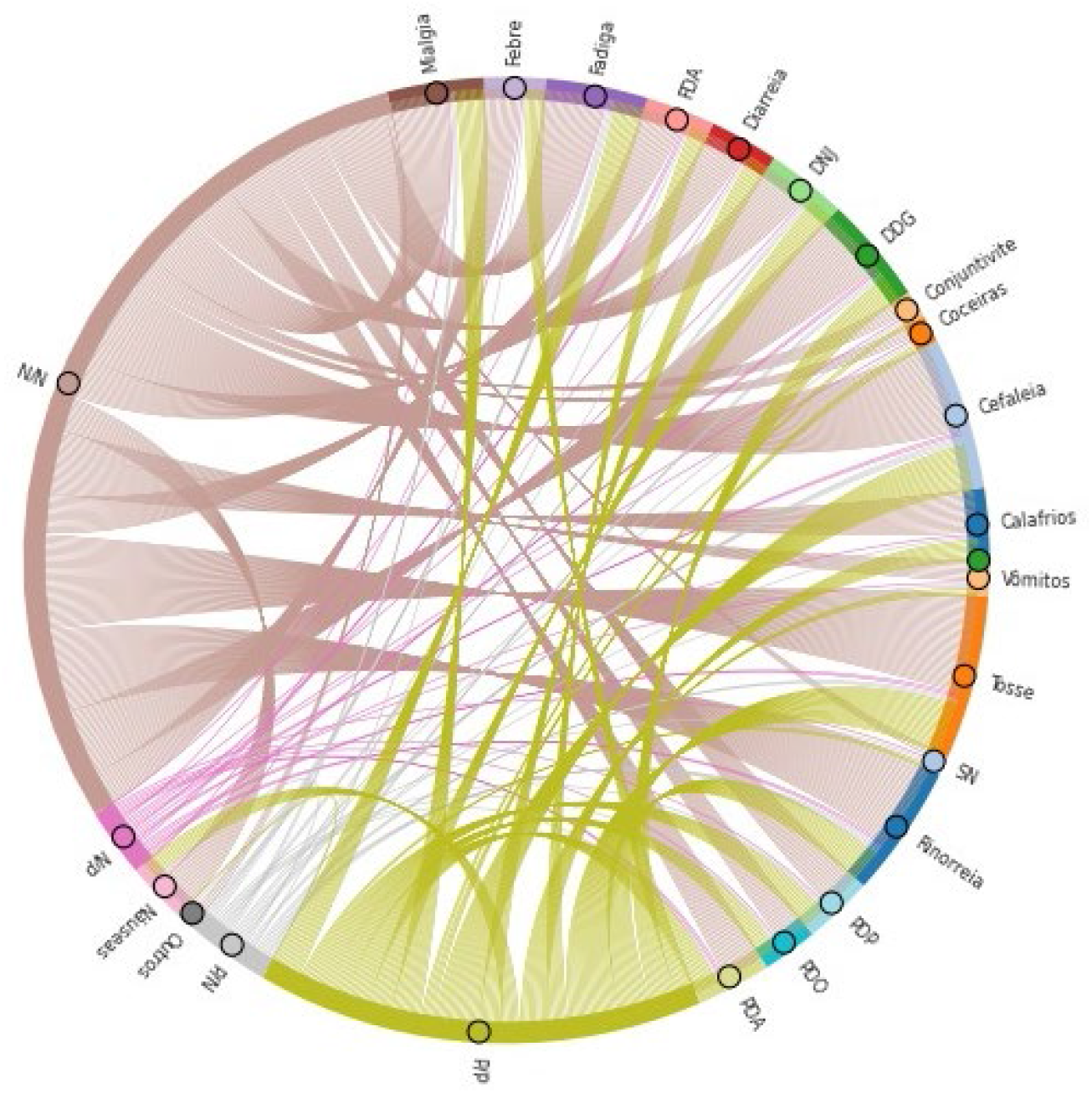
Symptoms correlated with positive and negative diagnoses of serological tests. Source: Developed by the Authors (2022)

It is possible to observe from Figure 5 that the share of positive results for detection of IgG and/or IgM antibodies is quite representative in the symptomatic picture of the research participants. This fact shows not only the increase incases of synd romes flu in this period of the year, but also the unpreparedness of government agents and the population in general regarding the emergence of a health crisis of global proportions and high infectious and lethal potential. Thus, it can be hypothesized those problems in the public health system, such as overcrowding in hospitals, were aggravated by the lack of preventive measures in the dissemination of the SARS-CoV-2 virus, which culminated in the precariousness of care services for those most in need.

Another factor to be found to be averse to efforts against the spread and lethality of new SARS-CoV-2 strains was the delay in the purchase and distribution of vaccines, a measure that would more efficiently mitigate the devastating effects on the Brazilian population.

It is of major importance to emphasize that the diagnosis of Covid-19 does not imply the exclusion of other concomitant diseases. A list of viral and bacterial agents that cause symptoms similar to those of Covid-19 is shown in Table 1.

**Table 1.** List of viral and bacterial agents causing symptoms similar to Covid-19.

| Viral agents |  |  | Bacteria |
| --- | --- | --- | --- |
| Adenovirus | Enterovirus | Viral parainfluenza | *Chlamydomphila pneumoniae* |
| Bocavirus (HboV) | Viral influenza | Rhinovirus | *Mycoplasma pneumoniae* |
| Coronavirus (except SARS-CoV-2) | Metapneumovirus | Respiratory syncytial virus (RSV) | *Mycobacterium tuberculosis* |

Source: Borghetti, A., Ciccullo, A., Visconti, E., Tamburrini, E., & Di Giambenedetto, S. (2020).

### 3.2 GENOME EVALUATION

As the pandemic holds high and rapid advance, there is a need to identify genetic risk factors, which can be fully harmful to humans due to their high capacity for mutation,hindering the possibilities of treatment and the creation of vaccines, evidencing the susceptibility of this serious disease. The genetic factors of the host, together with other risk factors, can help determine susceptibility to respiratory tract infections (13).

The purpose ofthis subsection is to bring analyses of the mutations that occurred in the SARS-CoV-2 samples collected in Brazil between June and November 2021, in relation to the original Wuhan strain. For this purpose, we used computational resources, as explained in the Methodology section.

The first analysis refers to the most frequent mutations in the strains analyzed. Thus, the 20 most frequent amino acid substitutions are presented in Figure 6 in descending order. The count of how many samples have taken acertain mutation is under the horizontal axis. The vertical axis indicates the gene and its mutation.

**Figure 6.**
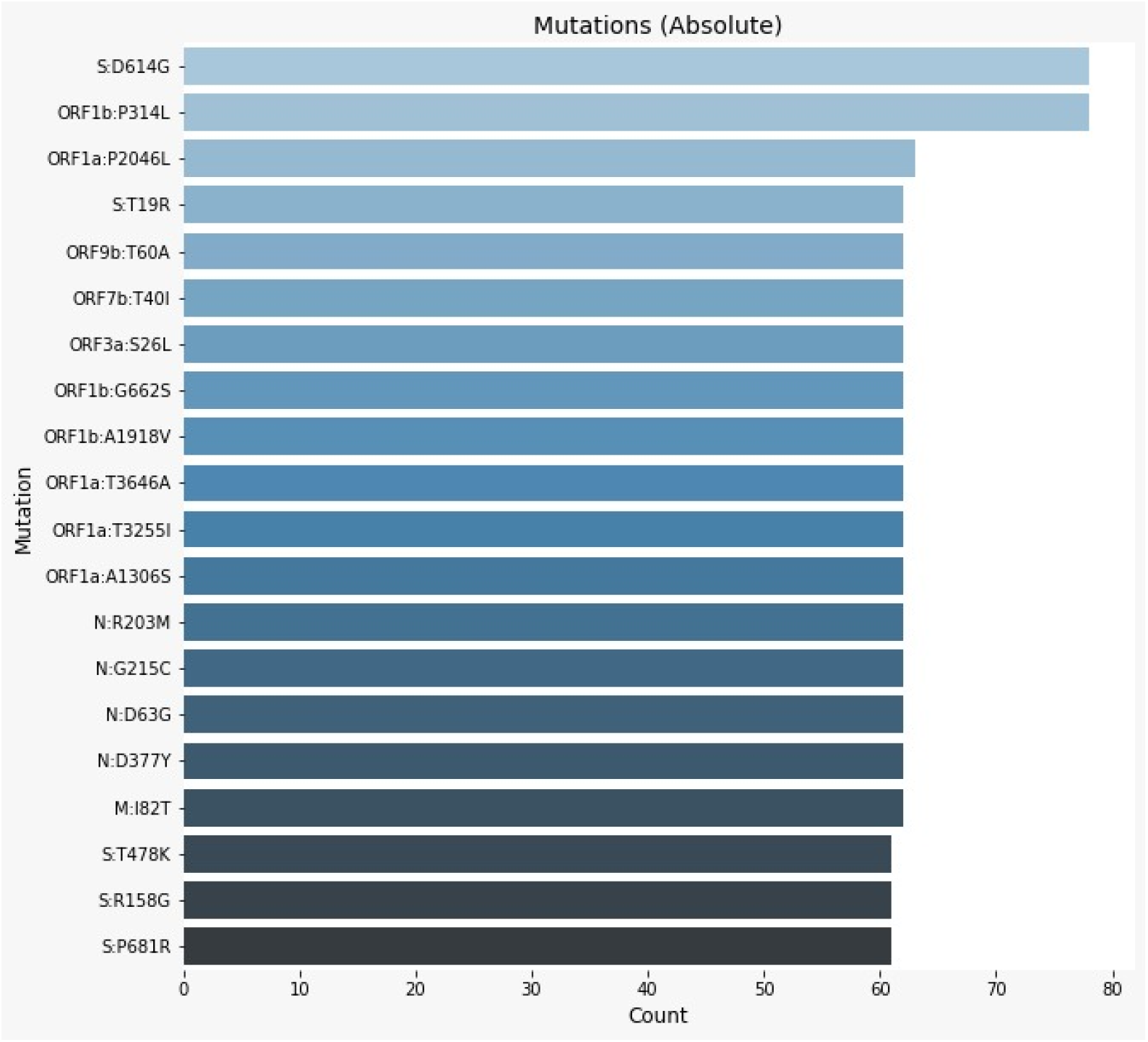
Most frequent amino acid substitutions in descending order. Source: Developed by the Authors (2022)

Following the results of other studies (14, 15), amino acid substitutions “S:D614G” and “ORF1b:P314L” are the most frequent, affecting approximately 100% of the analyzed samples. This indicates that themutations found in Brazilian strains of SARS-CoV-2 follow mutations found in strains from other parts of the world.

In relation to the mutation “S:D614G”, it occurs in position 614 of the spicule protein (S), where aspartic acid (D) is replaced by glylicina (G). Protein S is responsible for linking the virus to the host cell through the angiotensin-converter enzyme 2 (ECA 2) (14). Due to the function of protein S, studies indicate that the mutation “S:D614G” increases the transmissibility of the virus (14, 16), because it makes the interaction between the virus and ECA 2 more efficient (14). Although this mutation makes the virus more infectious, it does not seem to alter the severity of the disease (16).

The mutation “ORF1b:P314L”, also known as “NSP12:P323L” (14), occurs in an open reading phase (ORF) region, ORF1b, which is responsible, together with ORF1a, for synthesizing the 16 non-structural proteins (NSPs) of the new coronavirus (17). In relation to NSP, the mutation occurs in NSP12, also called RNA-dependent RNA polymerases (RdRps), which is responsible for the synthesis of viral RNA (15). Thus, the mutation occurs at position 323 of the 12th NSP, where proline (P) is replaced by leucine (L) (14).

Due to the high percentage of events of these two mutations and based on natural selection, it can be inferred that at least one of them actually contributes to the increased transmissibility of SARS-CoV-2.

The second analysis of genomic evaluation refers to the amount of amino acid substitutions in each of the coding regions of the SARS-CoV-2. The frequency of amino acid substitutions, in relation to the frequency expected at random, decreases linearly with the increase ofthe physical-chemical differences between the pairs of amino acids involved in a substitution, in a sense, the substitution frequencies can be dependent or independent of the physical-chemical parameters, the greater the difference between chemical properties, the greater the probability of amino acid substitution being attributed to natural selection (18).

In this analysis, only the first-time a given mutation occurred was counted; recurrences in other samples were not accounted for. The results are in Figure 7, where the x-axis represents the mutation count and the y-axis represents the genes.

**Figure 7.**
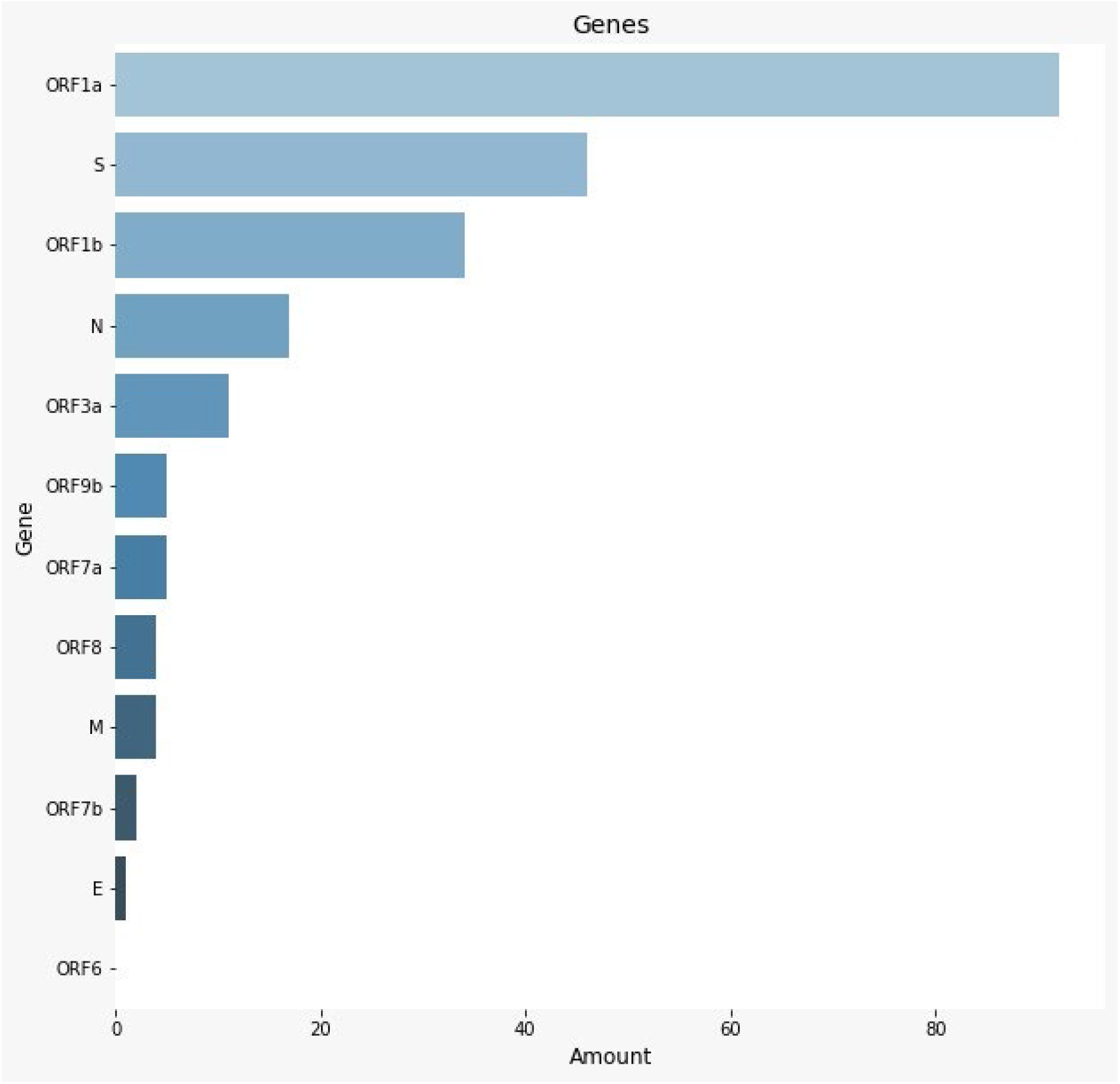
Number of amino acid substitutions in each of the coding regions of the SARS-CoV-2. Source: Developed by the Authors (2022)

The ORF1a gene is, with great difference, the gene with the most mutations. This is probably a consequence of the size of this coding region. ORF1a is the largest gene of SARS-CoV-2, containing 13202 nucleotides. In the analysis of the strains, with respect to quantity. The non-structural proteins that form the replication/transcription complex are encoded by genomic mRNA, where two overlapping ORFs are found. ORF1a encodes polyprotein 1a (pp1a) cleaved by viral proteases in each gene, detected: 11 non-structural proteins (nsp1– nsp11). The second (ORF1b) produces 5 additional nonstructural proteins (nsp12-16) (19).

The genome of the new coronavirus disease (Covid-19) was first sequenced in January 2020, approximately one month after its emergence in Wuhan, capital of Hubei Province, China. The sequencing of the COVID-19 genome is fundamental to understand the behavior of the virus, its origin, the speed with which it mutatesand for the development of drugs/vaccines and effective preventive strategies (20).

The entropy of genome information of the SARS-CoV-2 strains used in this study is shown in Figure 8. The axis of abscissa represents the 29903 nucleotides that make up the new coronavirus, in addition to the markings indicating the main coding regions of the virus. In sequence, from left to right, are: ORF1a, ORF1b, S, ORF3a, E, M, ORF6, ORF7a, ORF7b, ORF8, ORF9b and N. The axis of the ordered represents the entropy of the information. Here, the entropia refers to how mutated a given region is, so the higher the bar, the more amino acid substitutions happened in that region.

**Figure 8.**
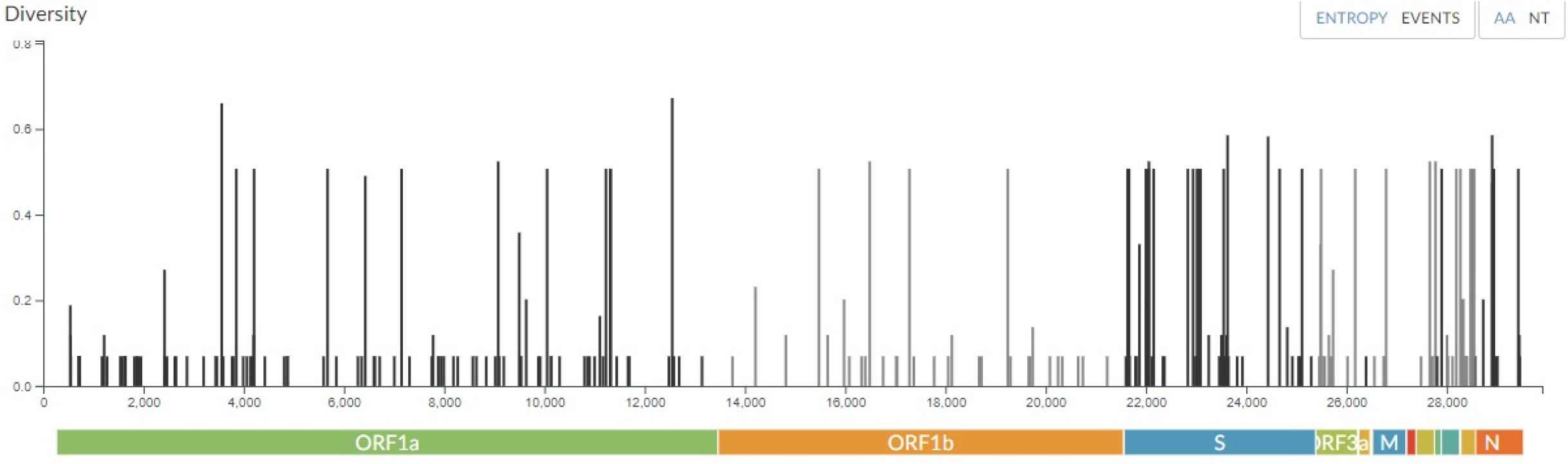
Genome Information Entropy.

As previously commented, ORF1a is the largest gene of SARS-CoV-2, and this can be noticed in the figure. In addition, a larger number of bars is visible in the ORF1a region, followed by S and ORF1b. The number of mutations is proportional to the number of bars, sincethis perception corroborates the previous result. When analyzing the entropy density in each of the genes, it is perceived that the S protein has the highest density, consequently, it is expected that this gene is the one that suffers the most mutations (21).

The analysis presented in Figure 9 shows the percentage of mutation in each coding region [the “percentage” will be in version 2 of the graph]. For this analysis, unlike the previous analysis, amino acid substitution recurrences were counted. The chart axes follow the same as in the previous analysis.

**Figure 9.**
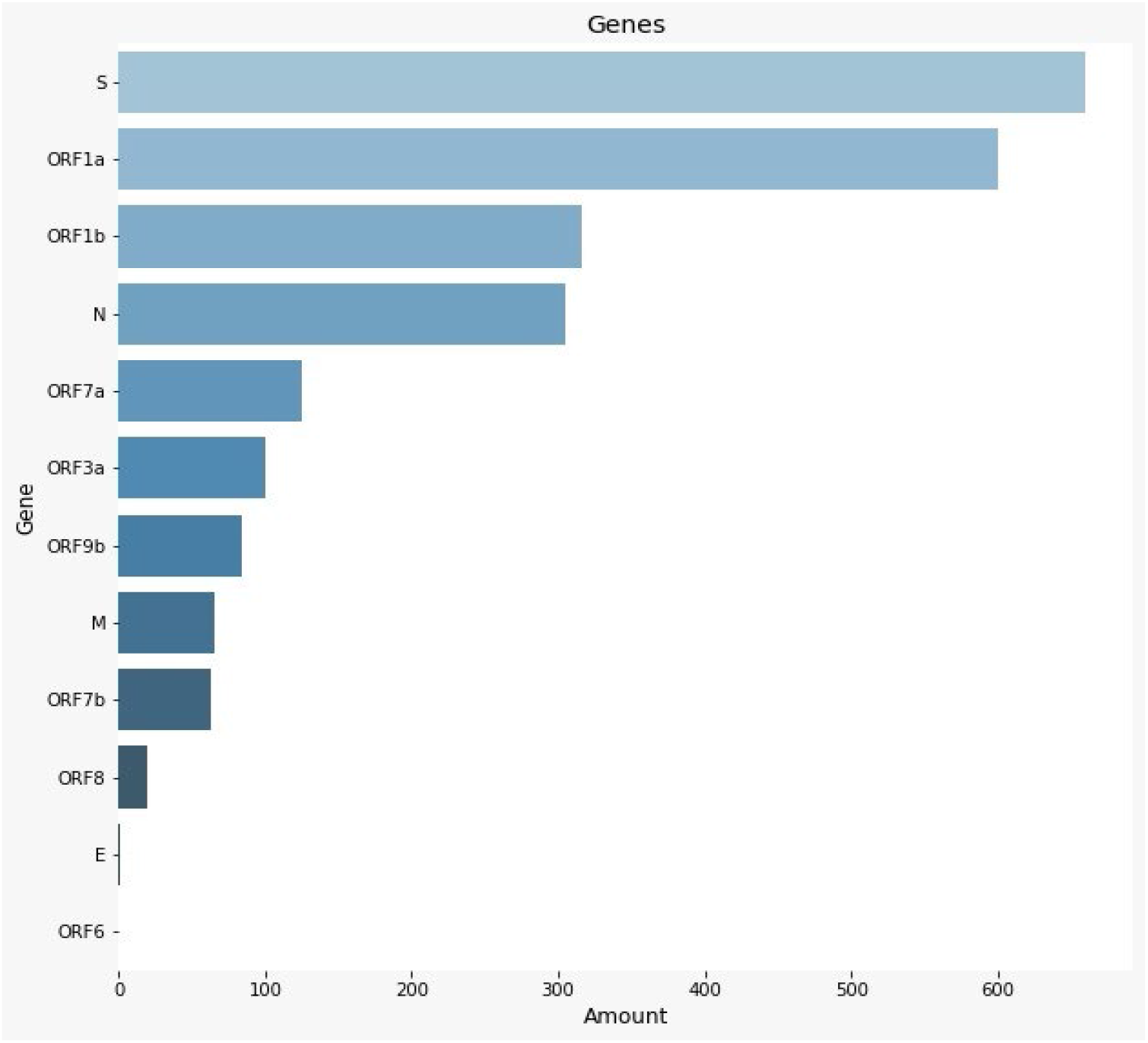
Percentage of mutation in each coding region. Source: Developed by the Authors (2022)

During the pandemic period, the Brazilian health system had full chronic problems of financing, management, provision of information and structuring of services in general. However, the lack of financial management and advances in denial, the variants were not fully controlled, causing greater mutations in Brazilian territory and diversified strains, even presenting one of the largest universal health systems in the world (22). In view of this, it is necessary tovaliate the Brazilian strains in phylogenetic tree system.

Figure 10 shows the phylogenetic tree of the Brazilian strains of SARS-CoV-2 between June and November 2021. The main elements of this phylogenetic tree are: the points representing the samples studied, the colors representing differentvariants of the virus, and the scale in the footer that represents the divergence of the sample in relation to the original Wuhan strain.

**Figure 10.**
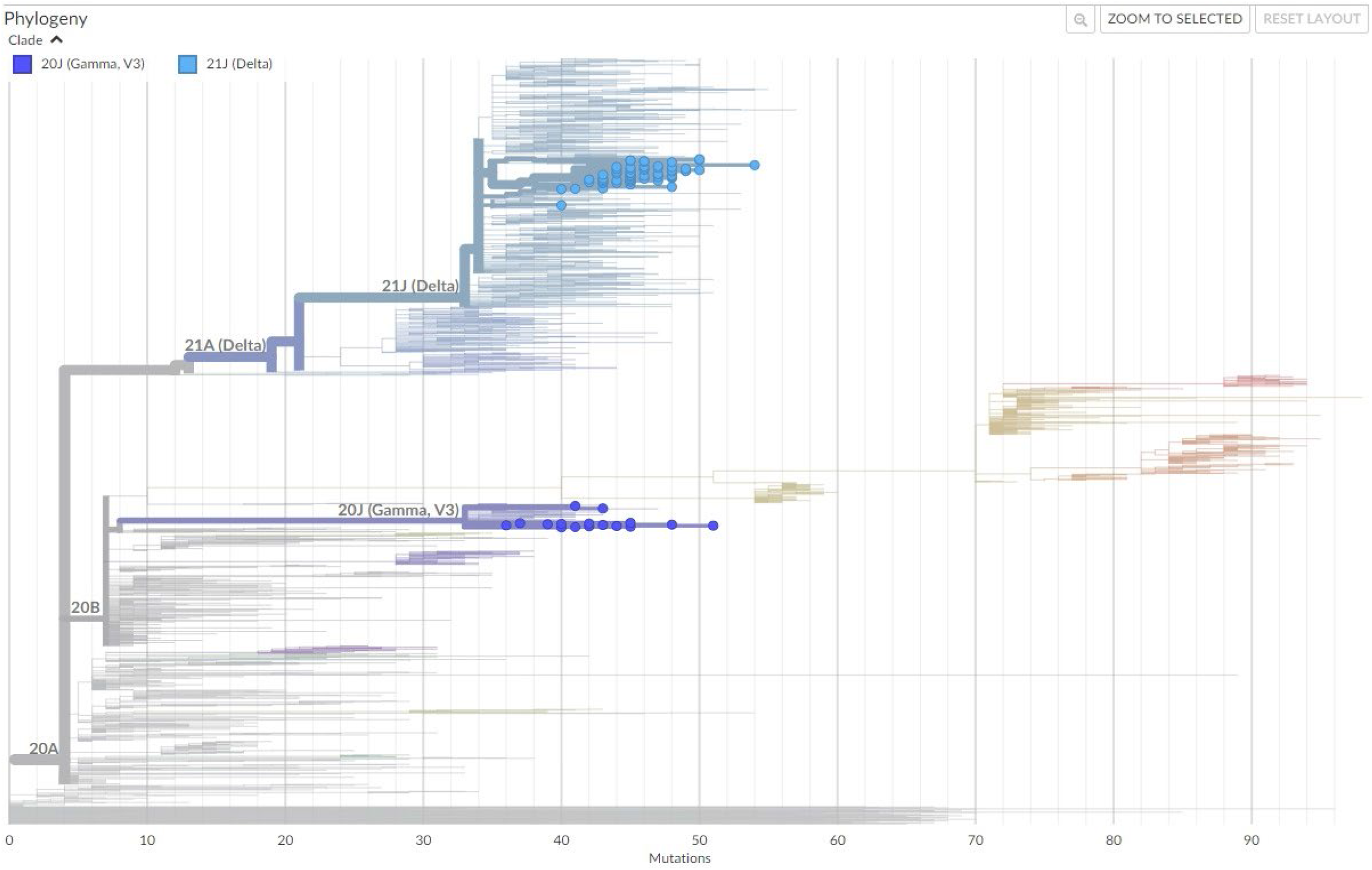
Phylogenetic tree. Source: Developed by the Authors (2022)

**Figure 11.**
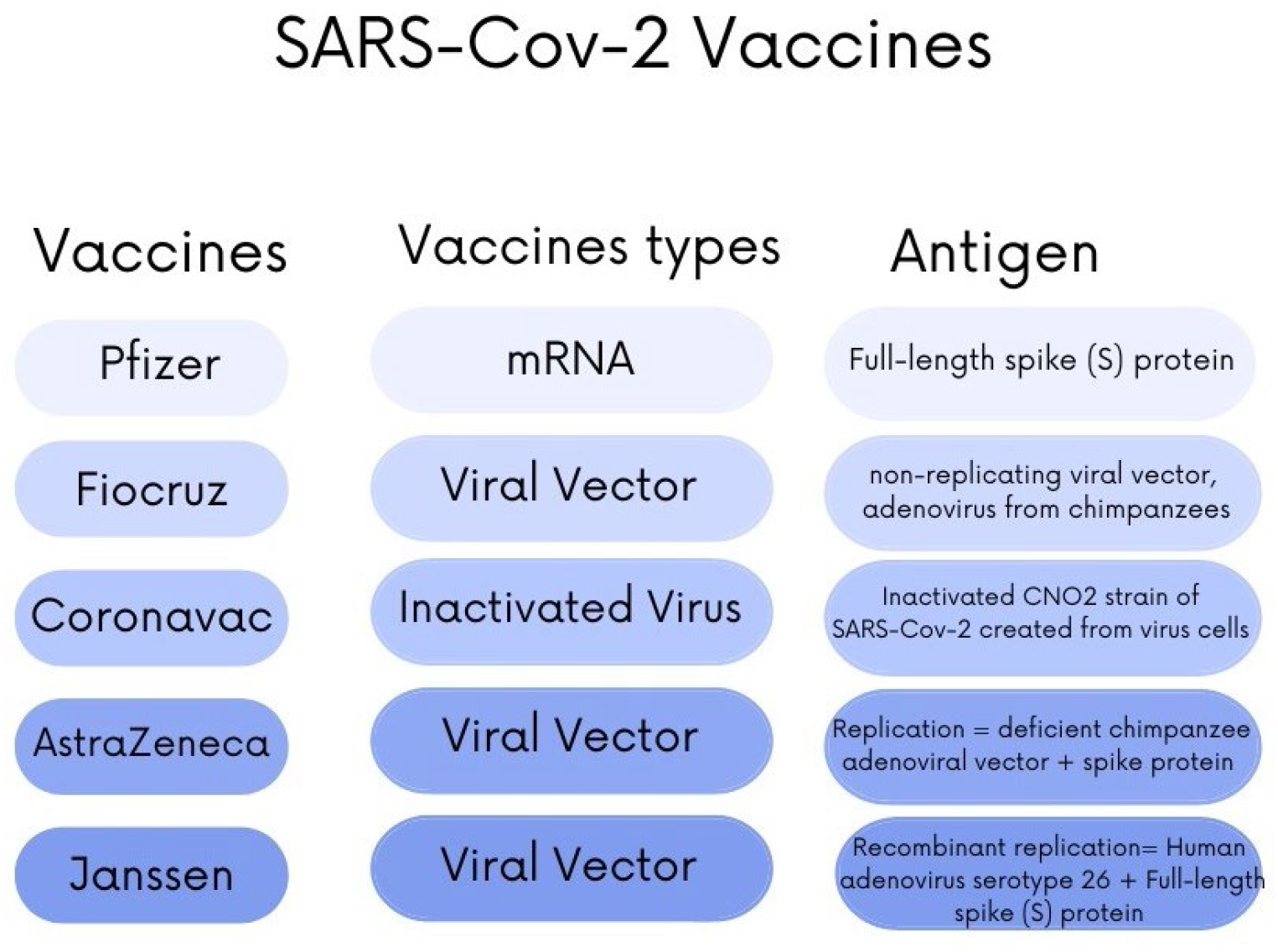
Vaccines, types of vaccines and antigens. source: Developed by the authors

Analyzing the phylogenetic tree, it is found that between June and November 2021, only delta and gamma variants circulated in Brazil. It is also noticed that despite the phylogenetic difference between these two variants, they have approximately the same divergence.

Several scientific articles already published corroborate the data provided by this article. Both the symptomatic question and the age group were declared in Brazil and outside it. According to Lima (23), in China, based on a study of 55,924 cases that were confirmed dand Covid-19, fever and cough were the most common symptoms, accompanied by fatigue, sputum production, dyspnea, sore throat, headache, myalgia, chills, nausea, nasal congestion, diarrhea, hemoptysis and conjunctive congestion, in this order. Monundo the same author (23), the first 99 patients hospitalized and evaluated in wuhan hospital, the highest rate came from over 50 years of age male.

Sousa Neto et al. (24) who focused on three pillars (adult people, symptomatic manifestations and Covid-19), also brought fever and normal or dry cough as symptoms associated with the disease, which was demonstrated in the present trial through graphs.

When it comes to the symptomatic age group of Covid-19, accordingto a study conducted in the state of R ondônia, Brazil, deaths occurred equally until 40 years of age, however, after this age, regardless of age group, the predominance of deaths came from males (25). Deaths for people over 60 years of age came from ord in52.7%, that is, more than half of the cases (25).

Finally, the study of the genome of Covid-19 was not behind when we talk about the importance of the present study, having knowledge of the types of variants and their genetic sequences may be the threshold between a rapid recovery or a large transmission, so the need to study which and how many mutations the Sars-Cov-2 gene is capable of. Michelon (26) considers that mutations in s protein are the most relevant when dealing from the clinical-epidemiological pointof view, due to its role in the infectious process, since it is a large transmembrane glycoprotein present on the external surface of Coronavirus. According to the author, the affinity of binding to the cell receptor characterizes higher transmission isssion, not different from that demonstrated in this article exposed by means of genomic mutations graphs of SARS-CoV-2.

### 3.3 IMMUNOLOGICAL FACTORS

SARS-CoV-2 is responsible for the development of acute respiratory syndrome, aggravated by symptoms: cough, headache and respiratory failures, which affect the entire oxygen absorption system. These symptoms are characteristic of the immune system process, essential to control and eliminate coronavirus infection (27).

The immune response of Covid-19 infection can be verified through rapid or serological tests. Serological tests for the diagnosis of Covid-19 are based on the linkingof antigens to antibodies, with different types of approaches, which can detect antibodies or viral antigens. The versions of commercial kits that detect antibodies against sars-cov-2 antigens that are most supplied on the market areof the s classes IgG, IgM or Total Antibodies (IgG + IgM), found in materials of total blood, serum or plasma samples of the patient (28).

However, the accuracy of serological tests varies by different factors, such as: methodology, antigen employed and time of collection (ideally between the 3rd and 10th days for IgM, IgA and total antibodies and, after 15th day, for IgG). In addition, other factors may lead to errors in test results, such as immunosenescence (state of immune function desgulated in the elderly that contributes to increased susceptibility to infections) (29), and the immunological window (period where serological tests are not able to detect the immune response) (30).

Moreover, both class M (IgM) and class G (IgG) immunoglobulin antibodies can be detected seven days after the onset of clinical symptoms, and may extend for more than 25 days, although the individual does not allow the individual to remain infectious, depending on his viral load and clinical presentation. The IgMvirus is 5 to 12 days after SARS-CoV-2 infection, followed by slow fall. IgG antibody reaches maximum concentrations after the 20th, approximately, as IgM antibodies disappear (31).

In addition, certain changes indiagnostics may also be correlated with the vaccines that had been administered in patients. Depending on the technology used, vaccines are manufactured in different approaches, causing different antibodies produced by the immune system.

An approach to vaccine development is the creation of inactivated vaccines derived from cultured viruses that are subsequently chemically inactivated, as evidenced in the Coronavac vaccine. In addition, another known approach is the vacinof viral vectors, which use viruses deficient in replication, designed to express the genetic sequence of the antigen of interest in host cells, such as Fiocruz. Janssen and AstraZeneca also use the same approach, but evidence of the spike *protein*. In addition, vaccines also use mRNA, in which lipid nanoparticles are used to protect the mRNA encoding the S protein stabilized by prefusion on the way to the intracellular space, as in Pfizer (32).

Depending on the technical approach used, the coding protein may not be identified in the tests, usually the S protein has a higher probability of identification, providing positive diagnosis. However, some vaccines do not have protein S and its constitution, that is, specific antibodies that would be identified in the IgG and IgM tests are not present in the individual’s body, but not necessarily it is without immunologic response, only stopped from relatively irrelev external factorsbefore, but that provided non-stimulation for the production of specific immunoglobulin. In a statement, this factor presented is also a possible impasse for the diagnosis of IgG and IgM serological tests (33, 34, 35).

Another determining agent for immunological factors are SARS-Cov-2 mutations and variants, which may compromise pre-established information by the immune system. After the emergence of the various waves of Variants of Covid-19, showing greater transmissibility and greater escape of antibodys (respectively, alpha, beta and delta), substantial escapes of the responses of neutralizing antibodies induced by vaccination or previous infection with a variant (36) were observed. In other words, the antibody titers did not have such an eficcia against the variants, as well as the serological tests, which demonstrated certain errors.

### 3.4 HUMANIST PERSPECTIVE

The notification rate of COVID-19 in Brazil was estimated at 9.2% (95% CI: 8.8% - 9.5%), and in all states, therates found were lower than 30%. São Paulo and Rio de Janeiro, the most populous states in the country, showed low notification rates (8.9% and 7.2%, respectively) (37). The Brazilian population, within the period, did not have full access to information and knowledge, notification rates were extremely low, compared to other countries.

Given that citizens have not been warned, the large information deficit is affirmable and salient, resulting in denialism, which translates into the acceptance of interventions without scientific validation, such as thedissemination and exaltation of unproven efficacy and efficacy with extremely serious side effects such as chloroquine, or the defense of an intervention strategy that contradicts the position of the World Health Organization (WHO), “vertical isolation”(38).

Moreover, the Brazilian government active in the period, promoted false information and dissemination of “*fake news*” since the initial pandemic period. The government failures statistically verified prove the increase in the number of deaths by Covid-19, which could have been avoided in a context of better administration (38).

The pandemic by Covid-19 is a public health emergency of international interest and represents one of the greatest challenges of humanity and science since World War II. The interface of this problem with the aspects of Mental Health and psychological resilience necessary for health professionals is also of fundamental importance, during and after the pandemic crisis (40). Active in the hospital area report such despair and suffering tothe neglectexperienced, however, the government again demonstrated itself powerless.

From the *One Health* perspective, the pandemic has had impacts on the triad, both in the animal health, environmental and primarily, human. Causing damage to the country directly, some irreversible (41).

## 4. CONCLUSION

The symptomatlogical analysis of the patients demonstrated the highest incidence of cough, ringororrhea and headache symptoms, as well as in relatively evident amounts of symptoms, being on average, reports of four, five or six symptoms together, salientando the symptomatic diversity that the SARS-Cov-2 virus can cause. In addition, the population profile can also be defined, specifying the age group: between 35 and 60 years.

The symptoms could be correlated with the results of serological tests, evidencing the symptomatic correlation with other infectious or inflammatory diseases and the positive and negative diagnoses of higher incidence for each symptom.

The genome of the virus was also evaluated, emphasizing the Brazilian strains in the period that the questionnaire was applied, between July and November 2021, a chaotic period in the pandemic context for Brazilian health. The genes with higher incidence, higher mutation and frequency were analyzed in a practical way, evidencing the coded spike proteins and ORFs. In addition, the strains were analyzed in a phylogenetic system in order to provide greater visualization of mutations applied to the specific time.

In short, the questionnaires applied provided relevant data, both for the historical context experienced, but for the changes in the social and political context. Symptomatic and genetic analyses are observable, directing the Brazilian context in a complete way, providing several interpretations and conclusions, both for the present and for future analyses.

## Data Availability

All data produced in the present study are available upon reasonable request to the authors. All data produced in the present work are contained in the manuscript.

